# Genetically predicted gene expression effects on changes in red blood cell and plasma polyunsaturated fatty acids

**DOI:** 10.1101/2024.12.17.24319171

**Authors:** Nikhil K. Khankari, Timothy Su, Qiuyin Cai, Lili Liu, Elizabeth A. Jasper, Jacklyn N. Hellwege, Harvey J. Murff, Martha J. Shrubsole, Jirong Long, Todd L. Edwards, Wei Zheng

## Abstract

Polyunsaturated fatty acids (PUFAs) including omega-3 and omega-6 are obtained from diet and can be measured objectively in plasma or red blood cells (RBCs) membrane biomarkers, representing different dietary exposure windows. *In vivo* conversion of omega-3 and omega-6 PUFAs from short-to long-chain counterparts occurs via a shared metabolic pathway involving fatty acid desaturases and elongase. This analysis leveraged genome-wide association study (GWAS) summary statistics for RBC and plasma PUFAs, along with expression quantitative trait loci (eQTL) to estimate tissue-specific genetically predicted gene expression effects for delta-5 desaturase (*FADS1*), delta-6 desaturase (*FADS2*), and elongase (*ELOVL2*) on changes in RBC and plasma biomarkers. Using colocalization, we identified shared variants associated with both increased gene expression and changes in RBC PUFA levels in relevant PUFA metabolism tissues (i.e., adipose, liver, muscle, and whole blood). We observed differences in RBC versus plasma PUFA levels for genetically predicted increase in *FADS1* and *FADS2* gene expression, primarily for omega-6 PUFAs linoleic acid (LA) and arachidonic acid (AA). The colocalization analysis identified rs102275 to be significantly associated with a 0.69% increase in total RBC membrane-bound LA levels (*P*=5.4×10^-12^). Future PUFA genetic studies examining long-term PUFA biomarkers are needed to confirm our results.

## INTRODUCTION

Polyunsaturated fatty acids (PUFAs) are primarily obtained from diet and include both short- and long-chain varieties of omega-3 and omega-6 fatty acids. It is known that eicosanoids generated from omega-6 metabolism may increase inflammation, whereas others derived from omega-3 metabolism are less inflammatory or anti-inflammatory. (Tapiero et al., 2002)

Prior to eicosanoid production, endogenous metabolism of omega-6 and omega-3 PUFAs occurs via a shared metabolic pathway including fatty acid desaturases [namely delta-5 desaturase (FADS1) and delta-6 desaturase (FADS2)], and elongase (ELOVL2). FADS2 and FADS1 are directly involved in converting long-chain PUFAs (i.e., linoleic acid, LA; and alpha-linolenic acid, ALA) into their respective shorter-chain varieties (i.e., arachidonic acid, AA; and eicosapentaenoic acid, EPA).(Marquardt et al., 2000) Whereas elongase is involved in the lengthening of the carbon chain to create long-chain omega-3 PUFAs, notably docosapentaenoic acid (DPA) and docosahexanoic acid (DHA).(Leonard et al., 2004)

Dietary intake in epidemiologic investigations can be measured using food frequency questionnaire requiring participant recall, or objectively assessed using a biomarker assay. For PUFAs, plasma biomarkers typically represent a shorter exposure window (approximately 24 hours) compared to PUFAs measured in red blood cell membranes (RBCs, or erythrocyte membranes) which usually approximate exposure windows over the past several months.(Arab, 2003) Given PUFA biomarkers, depending on how they were measured (either plasma or RBC), would represent different exposure windows, it is important to understand whether genetic variants associated with PUFA levels differ accordingly.

Several methods were employed in this analysis to comprehensively evaluate the role of genetics on PUFAs, measured in RBCs and plasma. To understand if potential differences between plasma versus RBC PUFA biomarkers are driven by gene expression, we compared genetically predicted gene expression (GPGE) on estimated changes in PUFA biomarkers at relevant metabolism genes (i.e., *FADS1, FADS2*, and *ELOVL2*). Furthermore, using colocalization, we sought to identify shared expression quantitative trait loci (eQTLs) associated with increased gene expression (at PUFA metabolism loci) and with changes in RBC PUFA biomarker levels.

## METHODS

### Polyunsaturated fatty acid GWAS

We conducted a GWAS of RBC fatty acids among polyp-free individuals of European ancestry controls (N=909) recruited in the Tennessee Colorectal Polyp Study (TCPS), a colonoscopy-based case-control study of adenomatous polyps conducted at Vanderbilt University Medical Center (VUMC) from 2003 to 2010 in Nashville, Tennessee. The GWAS for this analysis was conducted among polyp-free controls in order to prevent any concerns due to reverse causation of adenomas (a precursor lesion for colorectal cancer) potentially affecting measured levels of RBC PUFAs.(Amézaga et al., 2018) Details of the TCPS population and genetic data are provided elsewhere.(Edwards et al., 2013) Briefly, the source population for TCPS were patients between 45 and 70 years of age undergoing colonoscopy at the Vanderbilt Gastroenterology Clinic and the Veteran’s Affairs Tennessee Valley Health System in Nashville. Cases presented with polyps (i.e., adenomas, hyperplastic polyps, or both) and controls were polyp-free. Genotyping was conducted using Affymetrix Genome-Wide Human SNP Array 5.0 (Affymetrix, Inc.), and imputation was conducted using IMPUTEv2.2 using 1000 Genomes reference panels.(Edwards et al., 2013)

Fatty acid values were measured in red blood cells and represent a percentage of total membrane bound RBC phospholipid fatty acid content. Associations between genetic variants and the six RBC PUFAs (i.e., LA, AA, ALA, DPA, EPA, and DHA) were conducted using SNPTESTv2.5, assuming additive effect of each SNP allele on continuous change in RBC PUFA levels, and were adjusted for age, sex, and top 10 principal components for ancestry.

Publicly available GWAS data examining genetic associations with plasma PUFAs were also utilized in this analysis.(Guan et al., 2014; Lemaitre et al., 2011) These GWAS were conducted among individuals of European ancestry from the Cohorts for Heart and Aging Research in Genomic Epidemiology (CHARGE) consortium which includes 8,631 adults. PUFAs were measured in plasma and expressed as percentage of total fatty acids, and SNP associations with changes in plasma fatty acids were adjusted for age, sex, study site, and top principal components for ancestry.

### Selection of PUFA metabolism genes and relevant tissues

Metabolism of PUFA involves *in vivo* conversion of short-to long-chain omega-3 and omega-6 fatty acids via three genes, fatty acid desaturases (*FADS1, FADS2*), and elongase (*ELOVL2*).(Marquardt et al., 2000) Given their primary involvement in PUFA metabolism, the following three genomic regions were of interest in this analysis: the *FADS* region on chromosome 11 (i.e., *FADS1* GRCh37 bp 61317097-61584475, and *FADS2* GRCh37 bp 61583675-61884826); and *ELOLV2* region on chromosome 6 (GRCh37 bp 10730992-11294624). Furthermore, we focused the analysis on tissues most relevant for PUFA metabolism, namely adipose tissue,(Baylin et al., 2002) liver,(Frayn et al., 2006) muscle,(Frayn et al., 2006) and whole blood.(Hester et al., 2014)

### Genetically predicted gene expression (GPGE)

GWAS summary statistics from TCPS and CHARGE were utilized to estimated changes in RBC and plasma PUFA levels using S-PrediXcan. This approach uses an elastic net framework to estimate predicted changes in a specific trait driven by independent eQTL effects at a genomic locus.(Barbeira et al., 2018) S-PrediXcan leverages weights from gene expression prediction models in the Genotype-Tissue Expression (GTEx) project, covariances from a reference set (i.e., 1000 Genomes), and SNP-specific effects and standard errors from GWAS summary statistics for the selected traits (i.e., RBC and plasma PUFA GWAS for LA, AA, ALA, EPA, DPA, DHA). This approach results in tissue-specific changes in PUFA traits per standard deviation (SD) increase in gene expression, or genetically predicted gene expression (GPGE). For this analysis, S-PrediXcan was used to estimate GPGE effects for the three genes (i.e., *FADS1, FADS2*, and *ELOVL2*) and tissues relevant to PUFA metabolism (i.e., visceral adipose tissue, subcutaneous adipose tissue, liver, and whole blood). Summarized GPGE effects for each gene were estimated using random-effects meta-analysis and corresponding standard errors were estimated using the restricted maximum likelihood approach (REML) to account for possible heterogeneity of GPGE effects across tissues.

### Colocalization

For each genomic locus, colocalization was used to identify the putative causal variant for changes in RBC levels and compared to the GWAS hits for plasma PUFAs. The eQTL from GTEx in tissues relevant for PUFA metabolism were used in tandem with PUFA GWAS to identify shared variants involved with GWAS-significant changes in RBC and plasma PUFAs, and significantly involved in gene expression in PUFA metabolism tissues (i.e., visceral adipose tissue, subcutaneous adipose tissue, liver, skeletal muscle, and whole blood). Available information regarding GWAS variants, *P* values, minor allele frequencies, and PUFA effects (i.e., beta estimates) were leveraged in the colocalization procedure.(Giambartolomei et al., 2014)

## RESULTS

### GPGE for RBC PUFAs

In **Table 1**, we provide summarized REML random-effects meta-analysis effects for genetically predicted increased expression of *FADS1, FADS2*, and *ELOVL2* on changes in RBC PUFAs and plasma PUFAs in GTEx tissues with known involvement in PUFA metabolism. For RBC PUFAs, we observed increased expression of *FADS1* was associated with 2.75% increase in membrane-bound LA (*P*=0.31), 1.17% decrease in membrane-bound AA (*P*=0.25), and modest 0.16% and 0.21% increases in membrane-bound DPA and DHA, respectively. As shown in **Figure 1**, specific tissues potentially driving the REML random-effects summary for LA and AA at the *FADS1* locus included visceral adipose tissue (4.3% increase in LA, and 2.6% decrease in AA; **Supplementary Table 1**) and skeletal muscle (1.4% increase in LA, and 1.0% decrease in AA).

**Table 1.** Predicted changes in red blood cell (RBC) and plasma fatty acid levels associated with one standard deviation increase in genetically predicted gene expression (GPGE) for *FADS1, FADS2*, and *ELOVL2* genes summarized across GTEx tissues relevant to PUFA metabolism via random-effects meta-analysis.

| PUFAs | *FADS1* |  |  |  | *FADS2* |  |  |  | *ELOVL2* |  |  |  |
| --- | --- | --- | --- | --- | --- | --- | --- | --- | --- | --- | --- | --- |
|  | eQTL<sup>§</sup> | Beta | Std Err<sup>¶</sup> | *P* | eQTL<sup>§</sup> | Beta | Std Err<sup>¶</sup> | *P* | eQTL<sup>§</sup> | Beta | Std Err<sup>¶</sup> | *P* |
| **Red blood cell (RBC)<sup>†</sup>** |  |  |  |  |  |  |  |  |  |  |  |  |
| Linoleic acid (LA) | 36 | 2.747 | 1.458 | 0.31 | 87 | -1.025 | 0.310 | 4.5x10<sup>-02</sup> | 24 | -0.098 | 0.419 | 0.81 |
| Arachidonic acid (AA) | 36 | -1.168 | 0.475 | 0.25 | 87 | 1.550 | 0.543 | 0.07 | 24 | 0.908 | 0.782 | 0.25 |
| alpha-linolenic acid (ALA) | 36 | -0.004 | 0.008 | 0.72 | 87 | 0.007 | 0.005 | 0.21 | 24 | -0.002 | 0.010 | 0.86 |
| Eicosapentaenoic acid (EPA) | 36 | 0.028 | 0.060 | 0.72 | 87 | 0.060 | 0.027 | 0.11 | 24 | 0.148 | 0.078 | 0.06 |
| Docosapentaenoic acid (DPA) | 36 | 0.156 | 0.181 | 0.55 | 87 | 0.094 | 0.073 | 0.29 | 24 | 0.057 | 0.356 | 0.87 |
| Docosahexaenoic acid (DHA) | 36 | 0.205 | 0.212 | 0.51 | 87 | 0.394 | 0.037 | 0.09 | 24 | -0.066 | 0.292 | 0.82 |
| **Plasma<sup>‡</sup>** |  |  |  |  |  |  |  |  |  |  |  |  |
| Linoleic acid (LA) | 31 | -6.229 | 3.461 | 0.32 | 81 | 1.986 | 0.613 | 4.8x10<sup>-02</sup> | 24 | 0.035 | 0.045 | 0.43 |
| Arachidonic acid (AA) | 31 | 6.988 | 3.727 | 0.31 | 81 | -2.784 | 0.805 | 4.1x10<sup>-02</sup> | 24 | -0.167 | 0.084 | 4.9x10<sup>-02</sup> |
| alpha-linolenic acid (ALA) | 31 | -0.069 | 0.032 | 0.28 | 81 | 0.022 | 0.007 | 0.53 | 22 | 0.001 | 0.003 | 0.71 |
| Eicosapentaenoic acid (EPA) | 31 | 0.333 | 0.179 | 0.31 | 81 | -0.141 | 0.037 | 3.2x10<sup>-02</sup> | 22 | -0.084 | 0.019 | 1.43x10<sup>-05</sup> |
| Docosapentaenoic acid (DPA) | 30 | 0.305 | 0.156 | 0.30 | 81 | -0.134 | 0.037 | 3.6x10<sup>-02</sup> | 22 | -0.101 | 0.011 | 2.98x10<sup>-19</sup> |
| Docosahexaenoic acid (DHA) | 31 | 0.293 | 0.171 | 0.34 | 81 | -0.070 | 0.025 | 0.07 | 22 | 0.248 | 0.055 | 6.74x10<sup>-06</sup> |

<sup>†</sup>Expressed as % total membrane-bound fatty acids from the Tennessee Colorectal Polyp Study (TCPS) among polyp-free controls.
<sup>‡</sup>Expressed as % of total fatty acids from the Cohorts for Heart and Aging Research in Genomic Epidemiology (CHARGE) consortium.
<sup>§</sup>Expression quantitative trait loci (eQTL) included in the derivation of the GPGE in the following GTEx tissues with purported PUFA metabolism: adipose subcutaneous, adipose visceral, liver, skeletal muscle, and whole blood.
<sup>¶</sup>Standard errors for random-effects meta-analysis effects estimated using restricted maximum likelihood (REML).

**Figure 1.**
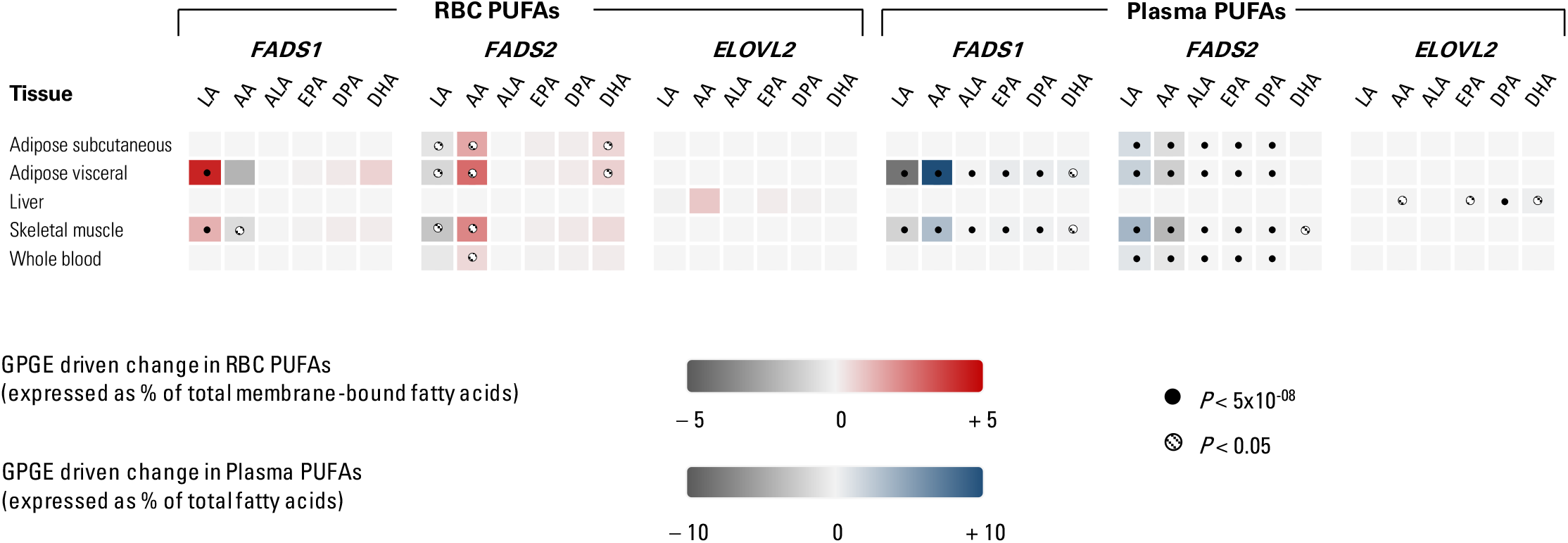
Genetically predicted gene expression (GPGE) for red blood cell (RBC) and plasma polyunsaturated fatty acids (PUFAs) in purported tissues involved in metabolism for increased *FADS1, FADS2*, and *ELOVL2* gene expression

In contrast, at the *FADS2* locus, increased gene expression was associated with a 1.03% decrease in membrane-bound LA (*P*=4.5×10^-02^), 1.55% increase in AA (*P*=0.07), and modest increases in membrane-bound DHA (0.39%). More tissues were involved in driving the GPGE at this locus, including adipose tissue (both visceral and subcutaneous), skeletal muscle and whole blood. The largest magnitude of increased RBC PUFAs was observed for increased AA in visceral adipose tissue (2.8%; **Supplementary Table 1**), and skeletal muscle (2.2%).

Finally, at the *ELOVL2* locus, modest overall changes in RBC PUFAs were observed with increased gene expression. A 0.91% increase in membrane-bound AA was suggested with one SD increase in *ELOVL2* gene expression (*P*=0.25; **Table 1**).

Furthermore, only liver tissue was involved in this observed increase in RBC PUFAs at this locus.

### GPGE for Plasma PUFAs

For plasma PUFAs (**Table 1**), one SD increase in *FADS1* expression resulted in 6.23% decrease in LA (of total plasma fatty acids), whereas a nearly 7.0% increase was indicated for AA. Modest increases on the accord of approximately 0.30% of total plasma fatty acids were observed for EPA, DPA, and DHA with increased *FADS1* expression.

Increased *FADS1* gene expression was predominantly observed in visceral adipose tissue and skeletal muscle tissue, where the largest magnitude of predicted change in plasma PUFA levels were observed for visceral adipose for LA (9.7% decrease; **Supplementary Table 1**) and AA (10.7% increase) for skeletal muscle. Similar patterns of predicted change in PUFA levels were also observed in skeletal muscle for LA (2.8% decrease) and AA (3.3% increase).

For *FADS2*, the largest magnitude of genetically predicted change across all tissues was observed for omega-6 PUFAs per one SD increase in gene expression, where LA (as percentage of total fatty acids) was observed to increase by nearly 2.0% (Std Err=0.61, *P*=4.8×10^-02^), and AA had a predicted 2.78% decrease (Std Err=0.81, *P*=4.1×10^-02^).

Modest changes in plasma EPA (-0.14%) and DPA (-0.13%) were observed with increased *FADS2* expression. Several PUFA metabolism tissues were identified (i.e., subcutaneous adipose tissue, visceral adipose tissue, skeletal muscle, and whole blood) where SD increases in gene expression was associated with changes in plasma PUFAs. The tissue with the largest magnitude of predicted increase in plasma LA was skeletal muscle (3.7%; **Supplementary Table 1**), followed by visceral adipose (2.1%), and subcutaneous adipose tissue (1.4%). On the contrary, total plasma AA was predicted to decrease by 4.9%, 3.1%, 1.9% at the same three tissues, respectively.

For increased *ELOVL2* expression, the magnitude for the predicted change in PUFAs were modest with AA and DHA having the largest magnitude of change with 0.17% decrease and 0.25% increase (of total plasma fatty acids), respectively. Several PUFA GPGE in liver tissue were statistically significant (i.e., plasma AA, EPA, DPA, and DHA). However, the magnitude for the predicted change for these plasma PUFAs was small ranging from -0.17% to 0.25% (**Supplementary Table 1**).

### Colocalization

A total of seven PUFA-specific top eQTLs located in *FADS1, FADS2*, and *ELOVL2* regions were associated with changes in RBC PUFA levels in plausible PUFA metabolism tissues (**Table 2**). The colocalization procedure identifies putative shared variants associated with both changes in RBC PUFA levels and gene expression, based on the predicted probability of this particular hypothesis.(Giambartolomei et al., 2014) One eQTL in the *FADS* locus (rs102275: Ch11:61557803) was identified in the colocalization analysis which was significantly associated with 0.69% increase in RBC LA levels, which explained approximately 5.4% in the variation in RBC LA levels. This variant was also associated with significant 1.46% decrease in plasma LA PUFA levels in previously published CHARGE GWAS.(Guan et al., 2014) In the *ELOVL2* region, rs210919 was suggested to decrease in RBC AA levels by approximately 1%; however, this variant was not significantly associated with changes in plasma AA. All other eQTLs identified to be associated with gene expression and changes in RBC PUFAs at the *FADS* and *ELOVL2* region explained less than or equal to 1% of the variation in the trait.

**Table 2.** Top expression quantitative trait loci (eQTL) effects on RBC PUFAs identified using colocalisation (posterior probability^†^ ≥0.90) limited to PUFA metabolism tissues in the Tennessee Colorectal Polyp Study (TCPS)

| PUFA | GRCh37 | eQTL | Effect/<br>Reference<br>allele | Effect<br>Allele<br>Frequency | Beta<sup>‡</sup> | Std Err | *P* | %<br>Variation<br>Explained |
| --- | --- | --- | --- | --- | --- | --- | --- | --- |
| ***FADS1* & *FADS2* region** |  |  |  |  |  |  |  |  |
| LA | 61557803 | rs102275 | T/C | 0.674 | 0.695 | 0.101 | 5.4x10<sup>-12</sup> | 5.42 |
| AA | 64835306 | rs492799 | C/A | 0.870 | -0.809 | 0.461 | 7.9x10<sup>-02</sup> | 0.38 |
| ALA | 61687454 | rs11230827 | G/A | 0.987 | 0.028 | 0.010 | 5.1x10<sup>-03</sup> | 0.95 |
| EPA | 62168194 | rs7127368 | G/A | 0.958 | 0.111 | 0.058 | 5.3x10<sup>-02</sup> | 0.46 |
| DHA | 61687454 | rs11230827 | G/A | 0.987 | 0.836 | 0.285 | 3.4x10<sup>-03</sup> | 1.06 |
| ***ELOVL2* region** |  |  |  |  |  |  |  |  |
| AA | 11710560 | rs210919 | G/A | 0.668 | -1.071 | 0.288 | 2.1x10<sup>-04</sup> | 1.64 |
| ALA | 11656361 | rs1150582 | A/G | 0.738 | -0.005 | 0.004 | 1.5x10<sup>-01</sup> | 0.26 |
| EPA | 11651138 | rs17399277 | G/T | 0.923 | -0.036 | 0.045 | 4.3x10<sup>-01</sup> | 0.08 |
| DPA | 11651138 | rs17399277 | G/T | 0.923 | -0.257 | 0.151 | 8.9x10<sup>-02</sup> | 0.36 |
| DHA | 11651138 | rs17399277 | G/T | 0.923 | -0.115 | 0.174 | 5.1x10<sup>-01</sup> | 0.05 |

<sup>†</sup>Colocalisation analysis posterior probabilities for shared variants associated with increased gene expression at PUFA metabolism loci (i.e., *FADS1*, *FADS2*, *ELOVL2*) in relevant PUFA metabolism tissues (i.e., adipose subcutaneous, adipose visceral, liver, skeletal muscle, and whole blood) and changes in RBC PUFA levels in TCPS.
<sup>‡</sup>Change in percentage of total membrane-bound RBC PUFA levels

## DISCUSSION

We sought to explore how genetically predicted gene expression for genes involved in PUFA metabolism in relevant tissues differed according to type of PUFA biomarker assayed. In general, we observed different magnitudes of effect at each genomic locus for changes in RBC versus plasma PUFAs. Increased *FADS1* gene expression was associated with increased genetically predicted RBC LA, whereas it was associated with decreased genetically predicted plasma LA. On the contrary, increased *FADS1* gene expression was associated with reduced RBC AA and increased plasma AA. Similarly, opposite changes in RBC versus plasma LA and AA were observed at the *FADS2* locus. Overall, only one eQTL was significantly associated with changes in LA measured in RBCs and increased gene expression at the *FADS* region.

Given *FADS1* plays a direct role in the conversion of short-chain LA to long-chain AA, the observed increased plasma AA levels seen with increased *FADS1* gene expression aligns with the biologic hypothesis. However, we did not observe the same pattern of increased long-chain AA with increased expression of *FADS2*, the other enzyme involved in the conversion. However, these results seen in plasma were opposite to what was observed in RBCs. It is possible that the GPGE results observed for RBC PUFAs reflects an important regulatory step in the production of long-chain PUFAs and eicosanoid production, where alleles in the same locus have opposing effects of downstream production of PUFAs.(Gieger et al., 2008; Mathias et al., 2010; Reynolds et al., 2020) One possibility is that increased gene expression of *FADS1* (the final enzymatic conversion to AA) only occurs when membrane-bound LA in RBCs is high, and increased *FADS2* gene expression occurs when membrane-bound LA levels is low. There is some evidence regarding the interaction between *FADS1* genotype with a diet high in LA on resulting PUFA concentrations, and possibility for feedback between fatty acid abundance and gene activity.(Meuronen et al., 2022; Tian et al., 2022) Our results indicate eQTLs involved in PUFA metabolism genes may have differential effects on objectively measured PUFA levels depending upon the type of biomarker measured (i.e., RBC versus plasma). Unfortunately, the magnitude of the results for omega-3 PUFAs (i.e., ALA, EPA, DPA, DHA) were less pronounced and this may be due to issues related to stability of the RBC biomarkers and possible degradation due to lipid peroxidation over time.(Metherel & Stark, 2016)

Several studies have identified environmental factors (primarily diet) that could influence the estimated effects of genetic variants on PUFA levels measured in RBC and plasma. Takkunen et al. examined the interaction between marine PUFA intake (assessed via food frequency questionnaire) with *FADS1* variant rs174550 on RBC PUFA levels and reported multiplicative interactions with dietary intake of EPA and DHA.(Takkunen et al., 2016) Al-Hilal et al. also reported significant changes in desaturase activity with long-chain PUFAs, post fish oil supplementation.(Al-Hilal et al., 2013) The previous studies examining changes in enzymatic activity were limited by small sample size. Nevertheless, the reported changes in activity are biologically plausible given corroboration in animal models where dietary intake of long-chain omega-3 fatty acids was reported to influence *FADS* enzymatic activity in several tissues (including muscle and adipose tissue).(Davidson et al., 2018)

Previous studies examining genetic influences on PUFA levels have primarily focused on eQTL effects on fatty acids measured in either RBC, plasma, or combined. A recent GWAS analysis identified variants associated with plasma PUFAs in Hispanic and African Americans, and reported SNPs residing in or near the *FADS* cluster and *ELOVL2* region.(Yang et al., 2023) The role of *FADS* region on changes in PUFA levels were also reported for RBC measured PUFAs in Chinese and European populations.(Hu et al., 2017) Previous studies that combined RBC measured PUFAs with plasma PUFAs as the outcome have also identified variants in the *FADS* region to play a role in increasing LA levels.(Hammouda et al., 2020; Mozaffarian et al., 2015) One study reported similar patterns of increased LA levels and reduced AA levels in RBC membranes associated with variants in the *FADS* cluster.(Rzehak et al., 2008) However, direct comparison to our analysis is difficult given the previous studies did not examine genetically predicted gene expression on changes in PUFA levels in relevant metabolism tissues.

Two-sample Mendelian randomization studies have become increasingly common over the years, and these studies rely upon existing genome-wide association study (GWAS) summary statistics to identify genetic instrumental variables for exposures of interest.(Haycock et al., 2016) Several studies have leveraged existing fatty acid GWAS to select genetic variants as instrumental variables for use in subsequent Mendelian randomization analyses with various outcomes. Given PUFA biomarkers may represent different exposure windows results from Mendelian randomization studies proxying nutrient biomarkers may also vary accordingly. The possibility for pleiotropic effects of genetic variants on other traits (including confounders) is inherent in existing GWAS summary statistics,(Bowden et al., 2017) thus increasing the specificity of the exposure contrast by selecting appropriate genetic proxies may help to further strengthen causal inference from these studies.(Swanson et al., 2017)

We identified differences in genetically predicted gene expression effects for PUFA metabolism genes according to PUFA biomarker (i.e., RBC versus plasma), which may be an important consideration for future studies on PUFAs utilizing genetic variants. Our study, while unique in its approach, is limited by the small TCPS sample size in which RBC PUFAs were measured. Furthermore, we used current eQTL information available in GTEx to estimate gene expression effects on PUFAs, which are limited by the number of tissues included in the resource. Future genetic studies of long-term PUFA biomarkers that are conducted in larger and more diverse populations are needed to confirm our results.

## Data Availability

All data produced in the present work are contained in the manuscript

## REFERENCES

Al-Hilal, M., AlSaleh, A., Maniou, Z., Lewis, F. J., Hall, W. L., Sanders, T. A. B., & O’Dell, S. D. (2013). Genetic variation at the FADS1-FADS2 gene locus influences delta-5 desaturase activity and LC-PUFA proportions after fish oil supplement. Journal of Lipid Research, 54(2), 542–551. 10.1194/jlr.P032276

Amézaga, J., Arranz, S., Urruticoechea, A., Ugartemendia, G., Larraioz, A., Louka, M., Uriarte, M., Ferreri, C., & Tueros, I. (2018). Altered Red Blood Cell Membrane Fatty Acid Profile in Cancer Patients. Nutrients, 10(12), 1853. 10.3390/nu10121853

Arab, L. (2003). Biomarkers of fat and fatty acid intake. The Journal of Nutrition, 133 Suppl 3(3), 925S–932S. 10.1093/jn/133.3.925S

Barbeira, A. N., Dickinson, S. P., Bonazzola, R., Zheng, J., Wheeler, H. E., Torres, J. M., Torstenson, E. S., Shah, K. P., Garcia, T., Edwards, T. L., Stahl, E. A., Huckins, L. M., GTEx Consortium, Nicolae, D. L., Cox, N. J., & Im, H. K. (2018). Exploring the phenotypic consequences of tissue specific gene expression variation inferred from GWAS summary statistics. Nature Communications, 9(1), 1825. 10.1038/s41467-018-03621-1

Baylin, A., Kabagambe, E. K., Siles, X., & Campos, H. (2002). Adipose tissue biomarkers of fatty acid intake. The American Journal of Clinical Nutrition, 76(4), 750–757. 10.1093/ajcn/76.4.750

Bowden, J., Del Greco M F., Minelli, C., Davey Smith, G., Sheehan, N., & Thompson, J. (2017). A framework for the investigation of pleiotropy in two-sample summary data Mendelian randomization. Statistics in Medicine, 36(11), 1783–1802. 10.1002/sim.7221

Davidson, E. A., Pickens, C. A., & Fenton, J. I. (2018). Increasing dietary EPA and DHA influence estimated fatty acid desaturase activity in systemic organs which is reflected in the red blood cell in mice. International Journal of Food Sciences and Nutrition, 69(2), 183–191. 10.1080/09637486.2017.1348494

Edwards, T. L., Shrubsole, M. J., Cai, Q., Li, G., Dai, Q., Rex, D. K., Ulbright, T. M., Fu, Z., Delahanty, R. H., Murff, H. J., Smalley, W., Ness, R. M., & Zheng, W. (2013). Genome-Wide Association Study Identifies Possible Genetic Risk Factors for Colorectal Adenomas. Cancer Epidemiology, Biomarkers & Prevention, 22(7), 1219–1226. 10.1158/1055-9965.EPI-12-1437

Frayn, K. N., Arner, P., & Yki-Järvinen, H. (2006). Fatty acid metabolism in adipose tissue, muscle and liver in health and disease. Essays in Biochemistry, 42, 89–103. 10.1042/bse0420089

Giambartolomei, C., Vukcevic, D., Schadt, E. E., Franke, L., Hingorani, A. D., Wallace, C., & Plagnol, V. (2014). Bayesian Test for Colocalisation between Pairs of Genetic Association Studies Using Summary Statistics. PLoS Genetics, 10(5), e1004383. 10.1371/journal.pgen.1004383

Gieger, C., Geistlinger, L., Altmaier, E., Hrabé De Angelis, M., Kronenberg, F., Meitinger, T., Mewes, H.-W., Wichmann, H.-E., Weinberger, K. M., Adamski, J., Illig, T., & Suhre, K. (2008). Genetics Meets Metabolomics: A Genome-Wide Association Study of Metabolite Profiles in Human Serum. PLoS Genetics, 4(11), e1000282. 10.1371/journal.pgen.1000282

Guan, W., Steffen, B. T., Lemaitre, R. N., Wu, J. H. Y., Tanaka, T., Manichaikul, A., Foy, M., Rich, S. S., Wang, L., Nettleton, J. A., Tang, W., Gu, X., Bandinelli, S., King, I. B., McKnight, B., Psaty, B. M., Siscovick, D., Djousse, L., Chen, Y.-D. I., … Steffen, L. M. (2014). Genome-wide association study of plasma N6 polyunsaturated fatty acids within the cohorts for heart and aging research in genomic epidemiology consortium. Circulation. Cardiovascular Genetics, 7(3), 321–331. 10.1161/CIRCGENETICS.113.000208

Hammouda, S., Ghzaiel, I., Khamlaoui, W., Hammami, S., Mhenni, S. Y., Samet, S., Hammami, M., & Zarrouk, A. (2020). Genetic variants in FADS1 and ELOVL2 increase level of arachidonic acid and the risk of Alzheimer’s disease in the Tunisian population. Prostaglandins, Leukotrienes and Essential Fatty Acids, 160, 102159. 10.1016/j.plefa.2020.102159

Haycock, P. C., Burgess, S., Wade, K. H., Bowden, J., Relton, C., & Davey Smith, G. (2016). Best (but oft-forgotten) practices: The design, analysis, and interpretation of Mendelian randomization studies. The American Journal of Clinical Nutrition, 103(4), 965–978. 10.3945/ajcn.115.118216

Hester, A. G., Murphy, R. C., Uhlson, C. J., Ivester, P., Lee, T. C., Sergeant, S., Miller, L. R., Howard, T. D., Mathias, R. A., & Chilton, F. H. (2014). Relationship between a common variant in the fatty acid desaturase (FADS) cluster and eicosanoid generation in humans. The Journal of Biological Chemistry, 289(32), 22482–22489. 10.1074/jbc.M114.579557

Hu, Y., Tanaka, T., Zhu, J., Guan, W., Wu, J. H. Y., Psaty, B. M., McKnight, B., King, I. B., Sun, Q., Richard, M., Manichaikul, A., Frazier-Wood, A. C., Kabagambe, E. K., Hopkins, P. N., Ordovas, J. M., Ferrucci, L., Bandinelli, S., Arnett, D. K., Chen, Y.-D. I., … Lin, X. (2017). Discovery and fine-mapping of loci associated with MUFAs through trans-ethnic meta-analysis in Chinese and European populations. Journal of Lipid Research, 58(5), 974–981. 10.1194/jlr.P071860

Lemaitre, R. N., Tanaka, T., Tang, W., Manichaikul, A., Foy, M., Kabagambe, E. K., Nettleton, J. A., King, I. B., Weng, L.-C., Bhattacharya, S., Bandinelli, S., Bis, J. C., Rich, S. S., Jacobs, D. R., Cherubini, A., McKnight, B., Liang, S., Gu, X., Rice, K., … Steffen, L. M. (2011). Genetic loci associated with plasma phospholipid n-3 fatty acids: A meta-analysis of genome-wide association studies from the CHARGE Consortium. PLoS Genetics, 7(7), e1002193. 10.1371/journal.pgen.1002193

Leonard, A. E., Pereira, S. L., Sprecher, H., & Huang, Y.-S. (2004). Elongation of long-chain fatty acids. Progress in Lipid Research, 43(1), 36–54. 10.1016/S0163-7827(03)00040-7

Marquardt, A., Stöhr, H., White, K., & Weber, B. H. F. (2000). cDNA Cloning, Genomic Structure, and Chromosomal Localization of Three Members of the Human Fatty Acid Desaturase Family. Genomics, 66(2), 175–183. 10.1006/geno.2000.6196

Mathias, R. A., Vergara, C., Gao, L., Rafaels, N., Hand, T., Campbell, M., Bickel, C., Ivester, P., Sergeant, S., Barnes, K. C., & Chilton, F. H. (2010). FADS genetic variants and ω-6 polyunsaturated fatty acid metabolism in a homogeneous island population. Journal of Lipid Research, 51(9), 2766–2774. 10.1194/jlr.M008359

Metherel, A. H., & Stark, K. D. (2016). The stability of blood fatty acids during storage and potential mechanisms of degradation: A review. Prostaglandins, Leukotrienes and Essential Fatty Acids, 104, 33–43. 10.1016/j.plefa.2015.12.003

Meuronen, T., Lankinen, M. A., Kärkkäinen, O., Laakso, M., Pihlajamäki, J., Hanhineva, K., & Schwab, U. (2022). FADS1 rs174550 genotype and high linoleic acid diet modify plasma PUFA phospholipids in a dietary intervention study. European Journal of Nutrition, 61(2), 1109–1120. 10.1007/s00394-021-02722-w

Mozaffarian, D., Kabagambe, E. K., Johnson, C. O., Lemaitre, R. N., Manichaikul, A., Sun, Q., Foy, M., Wang, L., Wiener, H., Irvin, M. R., Rich, S. S., Wu, H., Jensen, M. K., Chasman, D. I., Chu, A. Y., Fornage, M., Steffen, L., King, I. B., McKnight, B., … Arnett, D. K. (2015). Genetic loci associated with circulating phospholipid trans fatty acids: A meta-analysis of genome-wide association studies from the CHARGE Consortium. The American Journal of Clinical Nutrition, 101(2), 398–406. 10.3945/ajcn.114.094557

Reynolds, L. M., Dutta, R., Seeds, M. C., Lake, K. N., Hallmark, B., Mathias, R. A., Howard, T. D., & Chilton, F. H. (2020). FADS genetic and metabolomic analyses identify the Δ5 desaturase (FADS1) step as a critical control point in the formation of biologically important lipids. Scientific Reports, 10(1), 15873. 10.1038/s41598-020-71948-1

Rzehak, P., Heinrich, J., Klopp, N., Schaeffer, L., Hoff, S., Wolfram, G., Illig, T., & Linseisen, J. (2008). Evidence for an association between genetic variants of the fatty acid desaturase 1 fatty acid desaturase 2 ( FADS1 FADS2 ) gene cluster and the fatty acid composition of erythrocyte membranes. British Journal of Nutrition, 101(1), 20–26. 10.1017/S0007114508992564

Swanson, S. A., Tiemeier, H., Ikram, M. A., & Hernán, M. A. (2017). Nature as a Trialist?: Deconstructing the Analogy Between Mendelian Randomization and Randomized Trials. Epidemiology, 28(5), 653–659. 10.1097/EDE.0000000000000699

Takkunen, M. J., De Mello, V. D., Schwab, U. S., Kuusisto, J., Vaittinen, M., Ågren, J. J., Laakso, M., Pihlajamäki, J., & Uusitupa, M. I. J. (2016). Gene-diet interaction of a common FADS1 variant with marine polyunsaturated fatty acids for fatty acid composition in plasma and erythrocytes among men. Molecular Nutrition & Food Research, 60(2), 381–389. 10.1002/mnfr.201500594

Tapiero, H., Ba, G. N., Couvreur, P., & Tew, K. D. (2002). Polyunsaturated fatty acids (PUFA) and eicosanoids in human health and pathologies. Biomedicine & Pharmacotherapy = Biomedecine & Pharmacotherapie, 56(5), 215–222. 10.1016/s0753-3322(02)00193-2

Tian, H., Yu, H., Lin, Y., Li, Y., Xu, W., Chen, Y., Liu, G., & Xie, L. (2022). Association between FADS Gene Expression and Polyunsaturated Fatty Acids in Breast Milk. Nutrients, 14(3), 457. 10.3390/nu14030457

Yang, C., Veenstra, J., Bartz, T. M., Pahl, M. C., Hallmark, B., Chen, Y.-D. I., Westra, J., Steffen, L. M., Brown, C. D., Siscovick, D., Tsai, M. Y., Wood, A. C., Rich, S. S., Smith, C. E., O’Connor, T. D., Mozaffarian, D., Grant, S. F. A., Chilton, F. H., Tintle, N. L., … Manichaikul, A. (2023). Genome-wide association studies and fine-mapping identify genomic loci for n-3 and n-6 polyunsaturated fatty acids in Hispanic American and African American cohorts. Communications Biology, 6(1), 852. 10.1038/s42003-023-05219-w

