## Supplemental Table 1 for "Genetically predicted gene expression effects on changes in red blood cell and plasma polyunsaturated fatty acids"

**Supplementary Table 1.** Tissue-specific GPGE for RBC and plasma PUFAs

| PUFA | Tissue | Gene | RBC GPGE | RBC GPGE *P* | Plasma GPGE | Plasma GPGE *P* |
|---|---|---|---|---|---|---|
| LA | Liver | *ELOVL2* | -0.0983 | 8.14E-01 | 0.0353 | 4.32E-01 |
| LA | Adipose Visceral | *FADS1* | 4.2779 | 2.78E-10 | -9.6958 | 9.90E-258 |
| LA | Muscle Skeletal | *FADS1* | 1.3578 | 1.41E-10 | -2.7745 | 6.11E-203 |
| LA | Adipose Subcutaneous | *FADS2* | -0.7093 | 1.16E-02 | 1.3916 | 1.28E-29 |
| LA | Adipose Visceral | *FADS2* | -1.2242 | 9.43E-04 | 2.0658 | 1.23E-38 |
| LA | Muscle Skeletal | *FADS2* | -1.9313 | 7.28E-07 | 3.6738 | 3.95E-98 |
| LA | Whole Blood | *FADS2* | -0.5227 | 1.04E-06 | 0.8406 | 1.16E-67 |
| AA | Liver | *ELOVL2* | 0.9083 | 2.45E-01 | -0.1667 | 4.67E-02 |
| AA | Muscle Skeletal | *FADS1* | -1.0143 | 4.25E-02 | 10.7200 | 0.00E+00 |
| AA | Adipose Visceral | *FADS1* | -2.5867 | 8.91E-02 | 3.2652 | 1.10E-271 |
| AA | Adipose Visceral | *FADS2* | 2.7903 | 1.72E-03 | -1.9708 | 5.92E-142 |
| AA | Muscle Skeletal | *FADS2* | 2.2479 | 1.27E-02 | -3.1325 | 4.26E-205 |
| AA | Adipose Subcutaneous | *FADS2* | 1.5556 | 2.26E-02 | -4.8773 | 0.00E+00 |
| AA | Whole Blood | *FADS2* | 0.5433 | 2.66E-02 | -1.1655 | 6.65E-282 |
| ALA | Liver | *ELOVL2* | -0.0018 | 8.58E-01 | 0.0012 | 7.09E-01 |
| ALA | Muscle Skeletal | *FADS1* | -0.0041 | 6.21E-01 | -0.1014 | 1.41E-58 |
| ALA | Adipose Visceral | *FADS1* | 0.0014 | 9.64E-01 | -0.0368 | 1.10E-52 |
| ALA | Adipose Subcutaneous | *FADS2* | 0.0183 | 5.86E-02 | 0.0146 | 4.11E-08 |
| ALA | Adipose Visceral | *FADS2* | 0.0130 | 2.82E-01 | 0.0240 | 1.16E-11 |
| ALA | Muscle Skeletal | *FADS2* | 0.0104 | 3.99E-01 | 0.0422 | 2.59E-27 |
| ALA | Whole Blood | *FADS2* | 0.0021 | 5.22E-01 | 0.0096 | 1.94E-19 |
| EPA | Liver | *ELOVL2* | 0.1480 | 5.67E-02 | -0.0843 | 1.43E-05 |
| EPA | Muscle Skeletal | *FADS1* | 0.0258 | 6.86E-01 | 0.5132 | 2.64E-54 |
| EPA | Adipose Visceral | *FADS1* | 0.0465 | 7.99E-01 | 0.1556 | 1.91E-46 |
| EPA | Adipose Subcutaneous | *FADS2* | 0.1133 | 1.07E-01 | -0.1231 | 1.38E-17 |
| EPA | Whole Blood | *FADS2* | 0.0372 | 1.29E-01 | -0.1471 | 2.52E-16 |
| EPA | Muscle Skeletal | *FADS2* | 0.1196 | 1.73E-01 | -0.2382 | 7.71E-34 |
| EPA | Adipose Visceral | *FADS2* | 0.1035 | 2.28E-01 | -0.0596 | 9.05E-28 |
| DPA | Liver | *ELOVL2* | 0.0569 | 8.73E-01 | -0.1007 | 2.98E-19 |
| DPA | Muscle Skeletal | *FADS1* | 0.1485 | 4.37E-01 | 0.4619 | 7.34E-135 |
| DPA | Adipose Visceral | *FADS1* | 0.2215 | 6.90E-01 | 0.1498 | 1.12E-125 |
| DPA | Whole Blood | *FADS2* | 0.0709 | 3.93E-01 | -0.1012 | 7.28E-35 |
| DPA | Muscle Skeletal | *FADS2* | 0.2251 | 4.56E-01 | -0.1556 | 3.94E-47 |
| DPA | Adipose Subcutaneous | *FADS2* | 0.1363 | 5.32E-01 | -0.2271 | 1.72E-84 |
| DPA | Adipose Visceral | *FADS2* | 0.1806 | 5.44E-01 | -0.0550 | 1.76E-65 |
| DHA | Liver | *ELOVL2* | -0.0665 | 8.20E-01 | 0.2481 | 6.74E-06 |
| DHA | Muscle Skeletal | *FADS1* | 0.6706 | 2.99E-01 | 0.4765 | 1.00E-06 |
| DHA | Adipose Visceral | *FADS1* | 0.1495 | 5.05E-01 | 0.1342 | 3.01E-06 |
| DHA | Whole Blood | *FADS2* | 0.5963 | 2.66E-02 | -0.0780 | 9.84E-02 |
| DHA | Muscle Skeletal | *FADS2* | 0.7177 | 3.76E-02 | -0.0688 | 1.72E-01 |
| DHA | Adipose Subcutaneous | *FADS2* | 0.1543 | 1.02E-01 | -0.1684 | 4.08E-03 |
| DHA | Adipose Visceral Omentum | *FADS2* | 0.5118 | 1.32E-01 | -0.0383 | 1.88E-02 |
